# Estimating the reproductive number *R*_0_ of SARS-CoV-2 in the United States and eight European countries and implications for vaccination

**DOI:** 10.1101/2020.07.31.20166298

**Authors:** Ruian Ke, Steven Sanche, Ethan Romero-Severson, Nick Hengartner

## Abstract

SARS-CoV-2 rapidly spread from a regional outbreak to a global pandemic in just a few months. Global research efforts have focused on developing effective vaccines against SARS-CoV-2 and the disease it causes, COVID-19. However, some of the basic epidemiological parameters, such as the exponential epidemic growth rate and the basic reproductive number, *R*_0_, across geographic areas are still not well quantified. Here, we developed and fit a mathematical model to case and death count data collected from the United States and eight European countries during the early epidemic period before broad control measures were implemented. Results show that the early epidemic grew exponentially at rates between 0.19-0.29/day (epidemic doubling times between 2.4-3.6 days). We discuss the current estimates of the mean serial interval, and argue that existing evidence suggests that the interval is between 6-8 days in the absence of active isolation efforts. Using parameters consistent with this range, we estimated the median *R*_0_ value to be 5.8 (confidence interval: 4.7-7.3) in the United States and between 3.6 and 6.1 in the eight European countries. This translates to herd immunity thresholds needed to stop transmission to be between 73% and 84%. We further analyze how vaccination schedules depends on *R*_0_, the duration of vaccine-induced immunity to SARS-CoV-2, and show that individual-level heterogeneity in vaccine induced immunity can significantly affect vaccination schedules.

**Significance:** With the global efforts to develop vaccines for COVID-19, it is important to understand the contagiousness of the virus to design regional vaccination policy. To that end, we fit a mathematical model to data collected from the early epidemic period in the United States and eight European countries, estimating that the early epidemic doubles between 2.4-3.6 days. This suggests that SARS-CoV-2 is highly transmissible in the absence of strong control measures irrespective of heterogeneity in geographic and social settings. We estimated the median basic reproduction number, *R*_0_ to be 5.8 (confidence interval: 4.7-7.3) in the United States and between 3.6 and 6.1 in the eight European countries. The herd immunity needed to stop transmission is high (between 73% and 84%).

## Introduction

SARS-CoV-2 is the infectious agent that causes COVID-19. It is originated in Wuhan city, China in Dec, 2019 (1), and has spread rapidly causing the ongoing global pandemic. Initial estimates of the rate of early epidemic spread in Wuhan, China suggested that the epidemic grew at 0.1-0.14/day, leading to an epidemic doubling time of 5-7 days (2-5). However, using domestic travel data and two distinct approaches, we estimated that the epidemic in Wuhan grew much faster than initially estimated, and the growth rate is likely to be between 0.21-0.3/day before lock-down was implemented, translating to a doubling time between 2.3 and 3.3 days, and an *R*_0_ approximately at 5.7 with a confidence interval between 3.8 and 8.9 (6). However, it was not clear whether SARS-CoV-2 can spread in other geographic locations as fast as in Wuhan, China.

Accurate estimation of the rate of early epidemic growth is important for many practical aspects. First, it is crucial for forecasting the epidemic trajectory, the burden on health care systems (7, 8) and potential health and economic damage in the wake of potential second waves of COVID-19 outbreak (9). Second, it sets the baseline for evaluation of effectiveness of public health intervention strategies. Third, it is important for estimation of the basic reproductive number, *R*_0_, which in turn is used for predicting the herd immunity threshold needed to stop transmission (6, 10, 11). This is particularly pertinent to the development of vaccination strategies, especially given the current global effort to develop effective vaccines (12).

A major challenge to the inference of the growth of COVID-19 epidemic is that as a result of a fast-growing outbreak and a sizable infected population with no or mild-to-moderate symptoms (13, 14), case confirmation data is influenced by many factors in addition to the true epidemic growth, including substantial underreporting (15). Here, we argue that death and the cause of death are usually recorded more reliably and are less affected by surveillance intensity changes than case counts and that modeling the joint distribution of cases and deaths gives a more accurate assessment of COIVD-19 dynamics. It is possible that deaths from COVID-19 are underreported during very early phase of an epidemic when people are unaware of community transmission of COVID-19 (16), during the relatively late phase of the epidemic when health care system is overwhelmed (8), or for deaths in care homes. However, the underreporting is unlikely to significantly affect the growth rate of deaths as long as the rate of underreporting during the period between the early and the late phase of the epidemic remains roughly constant (17). We fit mechanistic models to both case and death count data collected from the United States (US) and eight European countries during this period, i.e. in early and mid-March, 2020 before broad interventions were established. We show that in most countries, COVID-19 spreads very rapidly, leading to high estimated *R*_0_ values and consequently high herd immunity thresholds in these countries. We further explore how vaccination schedules depend on the value of *R*_0_ and the distribution of the duration of vaccine-induced immunity in a population, in the context of the durations of protective immunity reported for SARS-CoV-2 (18, 19), as well as other coronaviruses such as HCoV-OC43 and HCoV-HKU1 (9, 20), SARS-CoV-1 (21-23) and MERS-CoV (24).

## Methods

### Data

We collected daily case confirmation and death count data for the US and eight countries in Europe from the John Hopkins CSSE (Center for Systems Science and Engineering) database (https://github.com/CSSEGISandData/COVID-19). The data is accessed and extracted on March 31, 2020. The data consists of time series of the cumulative number of case confirmations and deaths by country. Daily incidences were derived from the cumulative counts. We used data from the following countries: France (FR), Italy (IT), Spain (SP), Germany (GR), Belgium (BE), Switzerland (SW), Netherlands (NT), United Kingdom (UK) and the United States (US).

We included a subset of case and death count data for inference based on the two following criteria. First, to minimize the impact of stochasticity and uncertainty in early data collection, we used case confirmation incidence data starting from the date when the cumulative number of cases was greater than 100, and used daily new death count data starting from the date when the cumulative death count is greater than 20 in each country (see Table S1 and Fig. 1 for the period from which data is included). Second, to estimate the early outbreak growth in each country before control measures were implemented and at the same time maximize the power of inference, we allowed a maximum of 15 days of data points for the two types of data, leading to a maximum of 30 data points for each country. Note that the end date of incidence data used for inference is at or close to the date when strong control measures were implemented in each country (see Table S1), and thus the estimated growth rates represent the epidemic growth before broad measures were implemented. We also tested the sensitivity of model predictions when only 10, 13 days of data points are included for inference. The results are consistent across the different numbers of data points used (Table S1).

**Figure 1.**
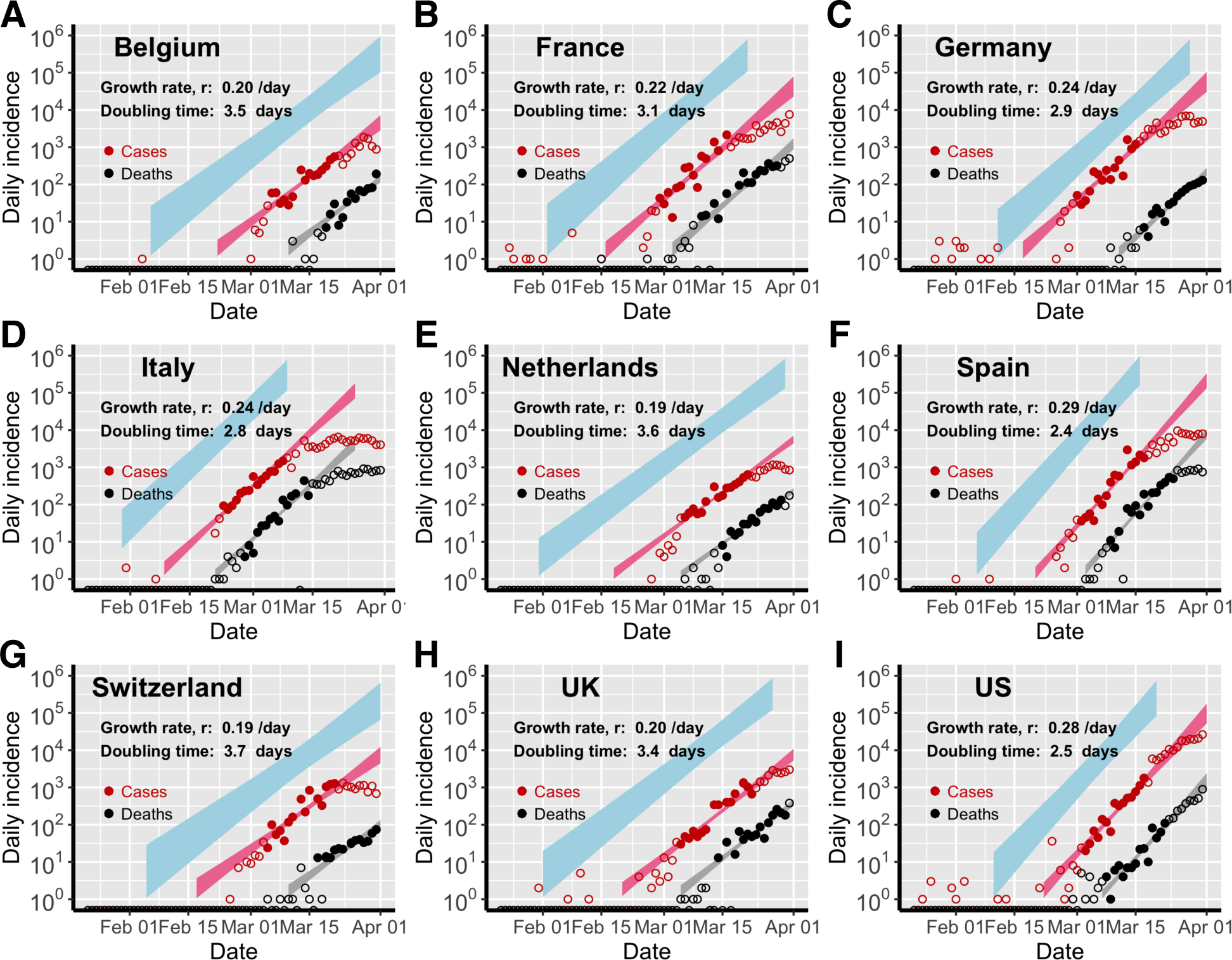
Estimation of the exponential growth rate and the doubling time of epidemics in eight European countries and the US. Red and black symbols show the daily counts of new cases and new deaths, respectively. Closed dots denote data used for parameter inference; whereas open circles denote data that are not used for parameter inference. We simulated the model using sampled parameter combinations that are able to explain the data shown in dots (see Uncertainty quantification in Methods). The colored bands denote the area between the lower and upper bounds of simulated/predicted true daily infection incidence (blue), daily cases (red) and daily deaths (grey) assuming no intervention efforts nor changes in surveillance intensity. Deviations of open circles from the corresponding bands thus indicate either changes in surveillance intensity or impacts of control measures.

### Model

We construct a SEIR type model using ordinary differential equations (ODEs; see Supplementary Text). We consider the exponentially growing phase of the outbreak and thus make the common assumption that the susceptible population is constant over time. Then, the total number of infected individuals *I*\*(*t*) = *E*(*t*) + *I*(*t*) can be expressed as:

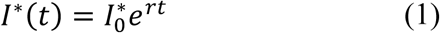

where *r* is the exponential growth rate of the epidemic (the growth rate for short below), and 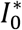 is the number of total infected individuals at time 0, set arbitrarily as January 20, 2020. Note the choice of the date of time 0 does not affect our estimation.

We solve the ODE model and derive the following expressions for the key quantities for model inference (see Supplementary Text). The descriptions and values used for the parameters in the ODE model are summarized in Table 1.

**Table 1.**
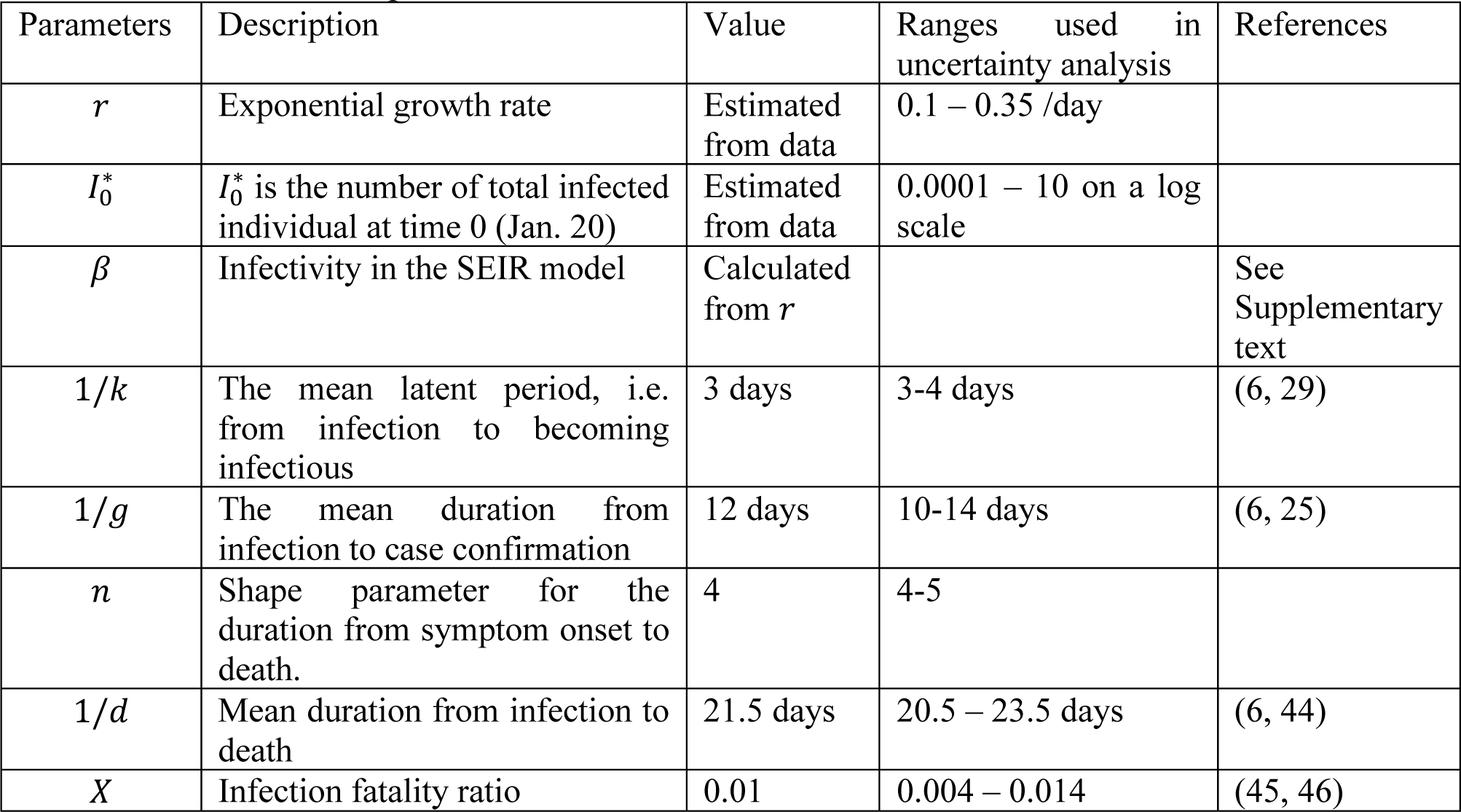
Description of parameter and their values. See the Supplementary Text for discussions of choice of parameter values.

| Parameters | Description | Value | Ranges used in uncertainty analysis | References |
| --- | --- | --- | --- | --- |
| $r$ | Exponential growth rate | Estimated from data | 0.1 – 0.35 /day | |
| $I_0^*$ | $I_0^*$ is the number of total infected individual at time 0 (Jan. 20) | Estimated from data | 0.0001 – 10 on a log scale | |
| $\beta$ | Infectivity in the SEIR model | Calculated from $r$ | | See Supplementary text |
| $1/k$ | The mean latent period, i.e. from infection to becoming infectious | 3 days | 3-4 days | (6, 29) |
| $1/g$ | The mean duration from infection to case confirmation | 12 days | 10-14 days | (6, 25) |
| $n$ | Shape parameter for the duration from symptom onset to death. | 4 | 4-5 | |
| $1/d$ | Mean duration from infection to death | 21.5 days | 20.5 – 23.5 days | (6, 44) |
| $X$ | Infection fatality ratio | 0.01 | 0.004 – 0.014 | (45, 46) |

The true daily incidence of infected individuals, Ω(*t*), can be expressed as:

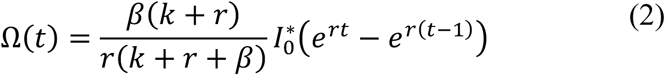

where β and 1/*k* are the transmission parameter of the virus and the latent period of infection, respectively.

The daily new confirmed case count, Ψ(*t*), is related to the true daily incidence, Ω(*t*) as:

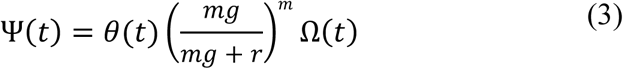

where *θ*(*t*) is the detection probability, i.e. the fraction of newly individuals at time *t* who are later detected among all infected individuals. We assumed an Erlang distribution for the period between infection and case confirmation (6), where 1/*g* and *m* are the mean and the shape parameter for the distribution.

The daily new death count, Φ(*t*), is related to the true daily incidence, Ω(*t*) as:

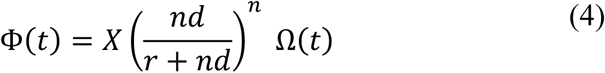

where X is the infection fatality ratio. Again, we assumed an Erlang distribution for the period between infection and death (6), where 1/*d* and *n* are the mean and the shape parameter for the distribution.

We tested three different scenarios for surveillance intensity changes over time, modeled as the detection probability, *θ*(*t*):

1. *θ* is a constant, i.e. no change over time;
2. 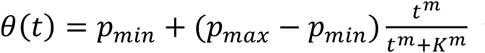 i.e. *θ* is a Hill-type function of *t*;
3. *θ*(*t*) is equal to *p*_*min*_ before *t*_1_, increases linearly to *p*_*max*_ between *t*_1_ and *t*_2_ and stay constant at *p*_*max*_ after *t*_2_, i.e. *θ* is a semi-linear function of *t*.

Note that the time from infection to case confirmation, 1/*g*, can be a time dependent function as we and others have shown previously (6, 25). To keep the model simple, we implicitly assume that the time dependent changes in *g* can be included in the estimation of *θ*(*t*).

### Parameter estimation

We fit the daily case count function Ψ(*t*) and the death count function Φ(*t*) to incidence data and daily death data to infer the exponential growth rate of the infection (*r*), the initial number of total infected individuals at time, and the detection probability (*θ*_*t*_). Other parameter values are fixed according to previous estimates (see Table 1). The error is calculated as the residual sum of squares (RSS) between data and model predictions of incidence or death counts on a log scale:

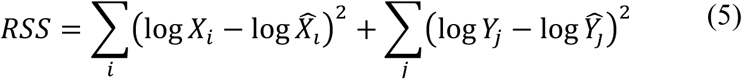

where *X*_*i*_ and 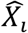 are simulated and reported new cases at day *i*, respectively, and *Y*_*i*_ and 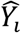 are simulated and reported new death cases at day *j*, respectively. Note that Eqn. 5 assumes log-normal error distributions of the case and the death counts. We estimate the parameter values by minimizing the RSS for each country.

To compare between models, we calculate the Akaike Information Criterion (AIC) score for each model as (26):

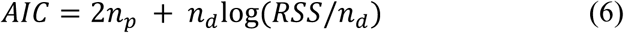

where *n*_*p*_ is the number of fitted parameters and *n*_*d*_ is the number of data points used in estimation. The model with the lowest AIC score is the best model. A model is significantly worse than the best model if the difference between their AIC scores is greater than 2 (26).

### Uncertainty quantification

We evaluate uncertainties in the estimated parameters r, *θ* and 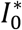, in two steps. First, we assess uncertainties in these parameters while keeping other parameters in the model fixed. We sampled 10^6^ parameter combinations of *r, θ* and 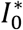 by drawing parameters randomly from uniform distributions over the ranges of the parameters r and 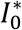 specified in Table 1. For the detection probability *θ*, we draw random numbers between 0,001 to 1 on a log scale. We calculate the RSS and the likelihood for each parameter combination. We accepted a parameter combination if the likelihood of this parameter combination was not statistically different from the best-fit parameter combinations using the log-likelihood ratio test. Second, to assess how estimation was impacted by uncertainties in the fixed parameters, we set different values for the mean latent period (2, 3 or 4 days), the mean time from infection to case confirmation (10, 12 or 14 days), the mean time from infection to death (16.5, 18,5 or 20.5 days) as well as the infection fatality ratio (0.4%, 0.8% or 1%). We then re-fit the model and assessed uncertainty in the estimated parameter values as described in the first step. The upper and lower bounds reported were summarized using simulation results of all accepted parameter combinations in the two steps.

### Estimation of *R*_0_

We calculated *R*_0_ according to the equation proposed by Wearing et al. (27):

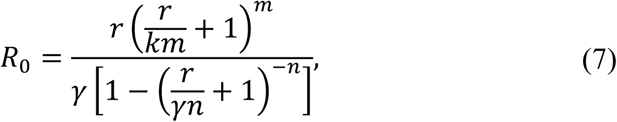

where 1/*k* and 1/*γ* are the mean latent and infectious periods, respectively, and *m* and *n* are the shape parameters for the gamma distributions for the latent and the infectious periods, respectively.

We set the mean latent period, 1/*k*, to vary between 3 and 4 days. This is based on that the incubation period is estimated to be between 5-6 days (6, 28, 29) and infected individuals become infectious approximately 2 days before symptom onset (30).

We set the mean infectious period, 1*/γ*, to be between 6-8 days to be consistent with the estimated mean serial interval, i.e. the mean time interval between symptom onsets of an index case and secondary cases in transmission pairs, of 6-8 days (3, 31, 32). See the Discussion section for a discussion of the estimates of the serial interval. We note that this range of infectious period is also consistent with the findings that infectious viruses can be recovered during the first week of symptom onset (and up to 9 days post symptom onset) (24, 33).

To quantify the uncertainty of *R*_0_, we assumed that m=4 and n=3 similar as in our previous work (6). We assume that the exponential growth rate, r, varies in the range estimated from the data. The parameters (*r, k, γ*) are assumed to be mutually independent and we generate random samples from uniform distributions according to ranges of variations defined above to compute the resulting *R*_0_. We generated 10^4^ parameters, and then computed their respective R_0_ using Eq. (7). We used the 97.5% and 2.5% percentile of the generated data to quantify the 95% confidence interval.

### Calculation of the level of population immunity after mass vaccination

We assume a gamma distribution for the duration of population immunity induced by a hypothetical vaccine to SARS-CoV-2. Let *τ* be the mean duration, and *s* be the shape parameter of the gamma distribution. For simplicity, we assume that the durations of the immunity induced by natural infection and vaccination are the same. We further assume that the percentage of protected population reaches to 85% after every mass vaccination with the hypothetical vaccine. Note that this is likely to be an optimistic scenario (34). The fraction of population that are immune to SARS-CoV-2 at time *t*^∗^ after a mass vaccination can then be expressed as 85% × (1 − C(*t*^∗^)), where C(*t*^∗^) is the cumulative density function of the gamma distribution for the duration of population immunity. Based on this expression, we calculate the time when the population immunity reaches to the herd immunity threshold value by solving 85% × -1 − C(*t*^∗^)) = 1 − 1/*R*_0_ for *t*^∗^. The solution for *t*^∗^ is the maximum time interval between two vaccinations to maintain herd immunity in a population.

## Results

### Estimation of the epidemic growth rate and surveillance intensity

Using our simplified SEIR-type (susceptible-exposed-infected-recovered) model (see Methods and Supplementary Text for details), we fit both the case incidence data and the daily death count data to estimate the epidemic growth rate and the detection probability, i.e. the probability that an infected person is identified, before interventions were implemented in eight European countries and the US. The exponential growth rates of early outbreaks, *r*, range between 0.19 and 0.29/day in the nine countries, translating to doubling times between 2.4-3.7 days (Fig. 1). Spain and the US had the highest estimated growth rates, at 0.29 and 0.28/day, respectively; whereas Switzerland and Netherlands had the lowest estimates at 0.19/day. Evaluating uncertainties in these estimates (see Methods), we found that the epidemic growth rates are highly constrained by the time series data despite variations in parameter values in the model (Fig. 2A).

**Figure 2.**
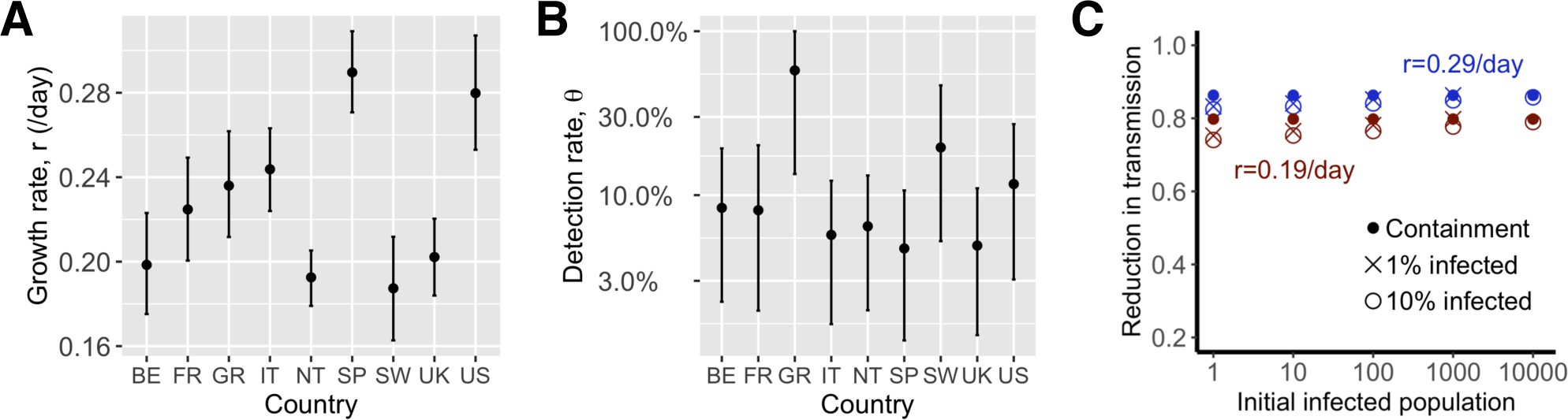
Fast spread of SARS-CoV-2 and its implications for public health interventions. Point estimates and confidence interval ranges of the exponential growth rate, *r* **(A)** and the detection probability, *θ* **(B)** in each country. See Table 2 for country name abbreations. **(C)** High levels of control efforts, measured as fractions of transmission reduction (y-axis), are needed to achieve containment, i.e. reverting epidemic growth (dots), or mitigation, i.e. the final fraction of infected individuals is 1% (x) or 10% (open circle) after a year. We assumed initial infected population as shown in x-axis and epidemic growth rates of 0.19 (red) or 0.29/day (blue).

We estimated that the detection probability, i.e. the fraction of infected individuals who are detected by surveillance, was likely to be low (<30%) across the countries examined except for Germany. The point estimate of the detection probability in the US is 12%, i.e. approximately 1 in 10 infected individuals were detected, similar to a recent estimate using influenza like illness data (35). This is likely due to the high percentage of infected individuals with no or mild-to-moderate symptoms (13, 14), which are difficult to detect through passive surveillance systems. The detection probability is higher in Germany (with a point estimate of 58%) than in other countries, providing an explanation of the high number of reported cases compared to the relative low number of deaths in Germany during March 2020. Overall, there exist large uncertainties in our estimation of the detection probability (Fig. 2B) due to the uncertainties in the fixed parameter values assumed in the model, such as the infection fatality ratio.

**Table 2.** **Estimated medians and confidence intervals of the basic reproductive number, R_0_, and their corresponding herd immunity thresholds for eight European countries and the US**.

| Country | Abbreviation | Median $R_0$ | Confidence interval - $R_0$ | Herd immunity threshold | Confidence interval – herd immunity |
| --- | --- | --- | --- | --- | --- |
| **Belgium** | BE | 3.8 | (3.2, 4.7) | 74% | (68%, 79%) |
| **France** | FR | 4.4 | (3.6, 5.4) | 77% | (72%, 81%) |
| **Germany** | GR | 4.7 | (3.8, 5.8) | 79% | (74%, 83%) |
| **Italy** | IT | 4.9 | (4.0, 5.9) | 79% | (75%, 83%) |
| **Netherlands** | NT | 3.7 | (3.2, 4.3) | 73% | (68%, 77%) |
| **Spain** | SP | 6.1 | (5.1, 7.5) | 84% | (80%, 87%) |
| **Switzerland** | SW | 3.6 | (3.0, 4.4) | 72% | (66%, 77%) |
| **United Kingdom** | UK | 3.9 | (3.3, 4.6) | 74% | (69%, 78%) |
| **United States** | US | 5.8 | (4.7, 7.3) | 83% | (79%, 86%) |

Changes to the detection probability over time, e.g. as a result of changes in testing, could lead to an apparent increase or decrease in case count and biases in inference. We considered two scenarios involving increases in testing over the study period (see Methods for the mathematical formulations), and found no statistical evidence that case counts during the relatively short period for which we perform inference are strongly impacted by changes in surveillance intensity (Table S2). While it is highly likely that the probability of a case being detected increased over the period where testing was becoming available, our analysis excluded data from that period. As shown in Fig. 1, the red, open circles indicate data outside of the study period. Most countries show a pattern of very rapid increase in detected cases in the very early epidemic period that is likely the result of both a growing epidemic and increasing availability and use of testing.

### Estimating the basic reproductive number, *R*_0_

We computed the basic reproductive number, *R*_0_, for each country following the approach of Wearing et al. (27), which uses as input the estimated growth rate, and the durations of the latent and infectious periods. We assumed that the duration of the latent period (i.e. the period between infection and becoming infectious) and the infectious periods to be 3-4 days and 6-8 days, respectively (see Methods for justification of these parameter ranges). These choices of parameters are consistent with the estimated mean serial intervals of 6-8 days (3, 31, 32). However, some estimates of the serial intervals are shorter (30, 36-38). In the Discussion, we present a more complete argument that for the purpose of estimating *R*_0_, a mean serial interval between 6-8 days is the current best estimate.

Using the estimated ranges of the growth rates for each country, we estimated that the US and Spain had highest median *R*_0_s at 5.8 (CI: 4.3-7.3) and 6.1 (5.1-7.5), respectively (Fig. 3 and Table 2). For the other countries, we estimated the median *R*_0_ ranges between 3.6 and 4.9 (Fig. 3 and Table 2).

**Figure 3.**
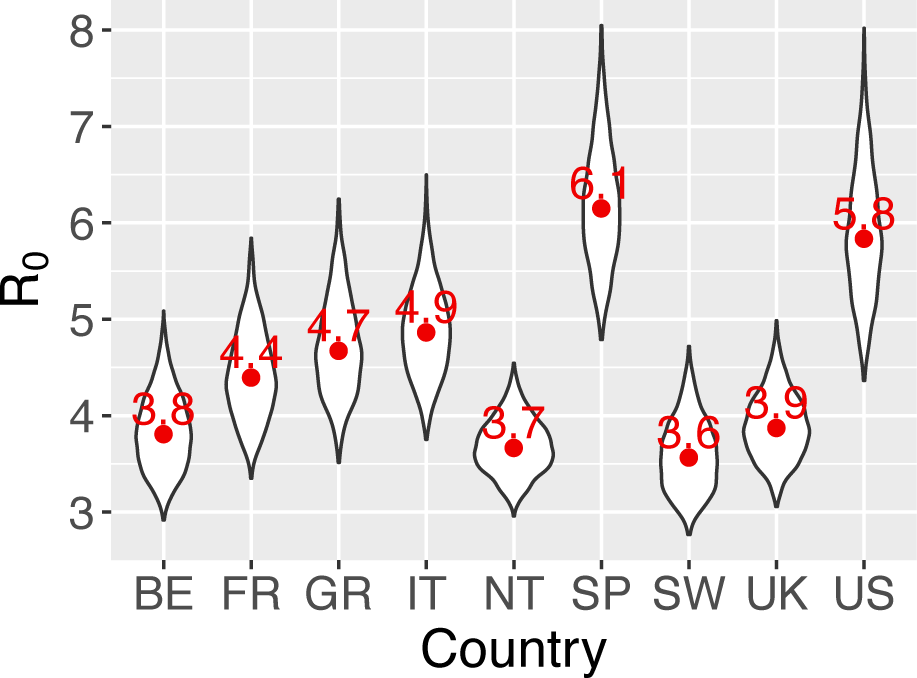
Point estimates and ranges of the reproductive number *R*_0_ in each country. Point estimates were shown in red numbers. See Table 2 for country name abbreations.

### Implications for public health intervention strategies

Using our empirical estimates of the growth rates, we explored the implications for public health efforts to control the COVID-19 outbreak. We considered an outbreak scenario in a large city with a population of 10 million, and three possible goals: 1) containment (i.e. the size of epidemic decreases), 2) 1% of the population is infected one year after the intervention, and 3) 10% of the population is infected one year after the intervention. Efforts needed for each goal are similarly high, especially when the population of infected individuals is already more than 1000 (Fig. 2C). For example, when an outbreak grows at rate 0.28/day (as we estimated for the US), the levels of efforts needed to achieve the three goals are between 81% and 84% reduction in transmission; whereas when the growth rate is 0.19/day, the levels of effort needed are between 71% and 75% reduction. Regardless of the heterogeneity in the growth rates, the force of infection must be significantly reduced, arguing for strong and comprehensive intervention efforts.

### Implications for vaccination strategies

From the range of *R*_0_ estimated above, we estimated that the range of herd immunity thresholds needed to prevent sustained transmission is between 73% and 84% using the formula 1-1/*R*_0_ (39) (Table 2). Multiple lines of evidence suggest that the protective immunity may not be long lived for SARS-CoV-2 (18, 19). Thus, we further considered how a vaccine with waning protection could be used to combat COVID-19 given our estimated levels of *R*_0_ (see Method). We assumed a gamma distribution for the duration of protective immunity induced by a hypothetical vaccine in a population, where *s* is the shape parameter of the gamma distribution (Fig. 4A).

**Figure 4.**
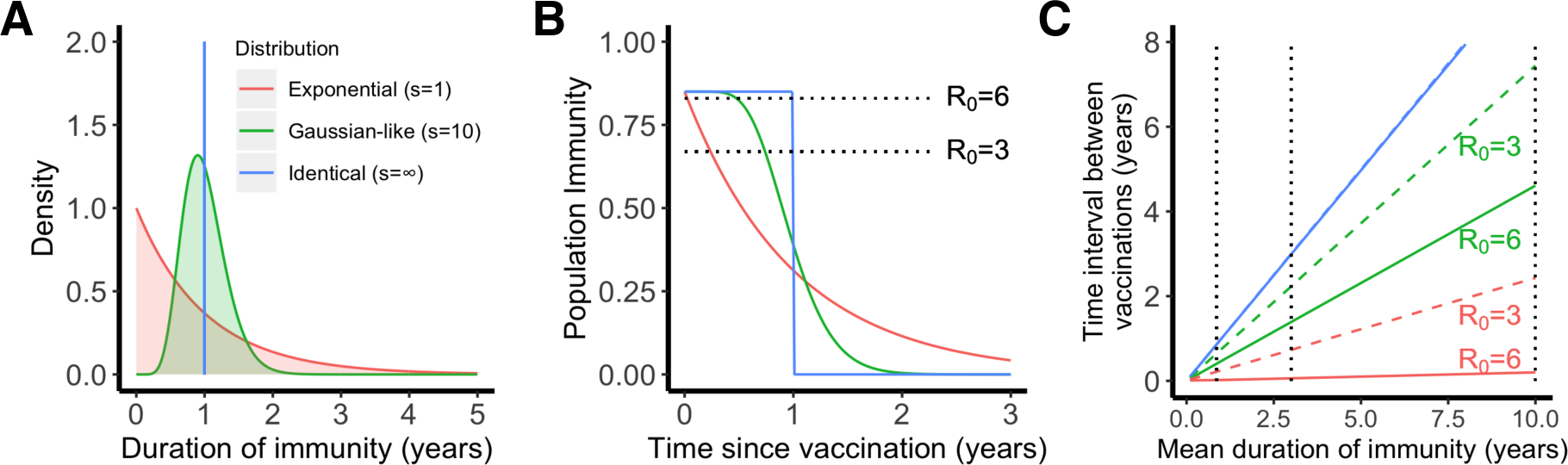
The importance of the distribution of the duration of vaccine-induced immunity in maintaining herd immunity in a population. **(A)** Three scenarios for the distribution: exponential (shape parameter *s* = 1), Gaussian-like (s = 10) and identical (s = ∞). All three distributions have the same mean, i.e. 1 year. The color code applies to all panels. **(B)** The fraction of individuals who are immune in a population over time. We assumed that 85% of population are immune after a mass vaccination at time 0. The dotted lines show the heard immunity thresholds, i.e. 83% (for *R*_0_=6) and 67% (for *R*_0_=3). **(C)** The time when the population immunity decreases to the threshold value (predicted for each *R*_0_) for the three scenarios. We assumed *R*_0_=3 (dashed lines) or 6 (solid lines) in the calculations. Note the dashed blue line overlaps with the solid blue line. The dotted lines show mean durations of immunity of 45 weeks, 3 years and 10 years.

If the duration of protective immunity from the vaccine follows an exponential distribution, (i.e. when *s* = 1), a sizable fraction of individuals lose immunity rapidly, leading to a loss of herd immunity shortly after the initial vaccination program, especially when *R*_0_ is large and the herd immunity threshold is high (Fig. 4B). Consequently, the time between vaccinations required to maintain herd immunity is much shorter than the mean duration of protective immunity (Fig. 4C). For example, even if the mean duration of protective immunity is 10 years, vaccination must occur every 2.4 month and 2.4 years to maintain herd immunity when *R*_0_ is 6 and 3, respectively (see the red lines in Fig. 4C). On the other extreme, when *s* is very large (Fig. 4A, *s* = ∞), individuals in the population have identical durations of protective immunity. In this case, herd immunity persists for a long period of time before the fraction of immune individuals suddenly drops to a very low level (Fig. 4B). In this case, herd immunity can be kept at a duration similar to the mean duration of protective immunity irrespective of *R*_0_ (see blue lines in Fig. 4C).

The reality of an imperfect vaccine is likely to be between these two extremes. When we assume *s* = 10, the distribution becomes more Gaussian-like (Fig. 4A) where some people lose protective immunity faster than others, but that heterogeneity is relatively low. If a mass vaccination achieves 85% immunity in a population and protective immunity lasts on-average around 45 weeks to 1 year (18) (consistent with the duration of immunity induced by endemic coronaviruses (9, 20)), then vaccination will need to occur once a few months (Fig. 4C). If the mean duration of immunity is around 3 years as observed for the antibody response to SARS-CoV-1 or MERS-CoV (24), vaccination once a year or once two years will be sufficient, if *R*_0_ is 6 or 3, respectively (Fig. 4C). If the mean duration of immunity is greater than 10 years (for example, a long T cell immunity to SARS-CoV-1 is observed in individuals recovered from SARS-CoV-1 infection (22)), the time interval between repeated vaccinations becomes longer than 4 years or 7 years when *R*_0_ is 6 or 3, respectively.

## Discussion

In this work, we report rapid COVID-19 epidemic spread before broad control measures were implemented in the eight most affected countries in Europe and in the US (during March, 2020). We further estimated that *R*_0_ values range between 3.6 and 6.1 in these countries, which means high herd immunity thresholds between 73% and 84%. Together with our previous estimates (6), these results are consistent with COVID-19 being highly transmissible in the absence of strong control measures irrespective of heterogeneities in geographic and social settings. A high level of coverage of effective vaccines are needed to achieve herd immunity. We further show that the heterogeneity of individual-level protection provided by a vaccine is an important factor in determining the frequency of vaccinations.

Awareness of the extraordinary high rates of COVID-19 spread is critically important for epidemic preparedness, especially in face of the potential second wave of infection in the fall of 2020 (9). Regardless of the country, the short doubling times we estimated means that health care systems can be overwhelmed in a couple of weeks rather than several months in the absence of control (8). For example, a report shows that the number of COVID-19 patients admitted to intensive care units in Italy grew at a rate of approximately 0.25/day during early epidemic (7). While we found remarkably high levels of spread in all examined countries, we caution that our inference is largely driven by data collected from highly populated areas, such as Wuhan in China, Lombardy in Italy, and New York city in the US. Heterogeneities in the growth rate almost certainly exist among different areas within each country. For example, recent works suggest that the rate of spread is positively associated with population densities (40). Therefore, the estimates we provide may represent good estimates in populated areas such as in cities.

Calculation of the basic reproductive number requires knowledge of the duration and distribution of the serial interval, which in turn is determined by the latent and the infectious periods. By explicitly considering the distributions of the latent and the infectious periods, our work more accurately incorporate the timing of transmission events in the estimation of *R*_0_ (27). Our estimates of *R*_0_ for several European countries are slightly higher than the estimated effective reproductive numbers during early dynamics reported in a work published recently (17). The explicit consideration of the distributions of the latent and the infectious periods and a slightly longer serial interval assumed in our approach are likely to be the cause of this discrepancy.

Various estimates of the mean serial interval (SI) exist for COVID-19. Similar to our earlier work (6), we assumed parameter values that are consistent with a mean SI of 6-8 days. This is based on estimates using data from Wuhan, China and Vo, Italy (3, 32). Shorter mean SIs were reported in the literature, for example, 4.0 days in Du *et al*. (36), 5.8 days in He *et al*. (30), 4-5 days in Nishiura *et al*. (37), 4-5.2 days in Ganyani *et al*. (38). However, these estimates were based on transmission pairs reported in locations outside of Wuhan, Hubei province, and other Asian countries and territories neighboring China, where active surveillance and rapid isolation programs were implemented. Indeed, we previously estimated that in provinces outside of Hubei province, the mean time from symptom onset to hospitalization/isolation, was as short as 1.5 days after Jan 18^th^. A short duration between symptom onset and hospitalization was also estimated for the outbreak in Singapore (25). This is in stark contrast to the time to hospitalization estimated for patients in Wuhan (9-13 days (3)). Rapid isolation of infectious individuals prevents further transmission after isolation, and thus inference of SIs from the transmission pairs that were rapidly isolated will be biased towards shorter values. The impacts of active tracing and isolation have been demonstrated by Bi *et al*. (31) and discussed in Ref. (41). Bi *et al*. showed that in transmission pairs where index cases were isolated within 3 days after symptom onset, the estimated mean SI is 3.6 days on average; whereas when index cases were isolated after 3 days post symptom onset, the estimated mean SI becomes much longer, approximately 8.0 days. Importantly, a recent work estimated the mean serial interval based on data from and Vo, Italy (32), and showed that the mean SI is 7.2 days, consistent with estimates from Wuhan, where little activate isolation efforts were in place during early epidemic (3). Therefore, to calculate *R*_0_ (in the eight European countries and the US where minimal active tracing and isolation programs were implemented), a serial interval between 6-8 as estimated in Refs. (3, 31, 32) represents the best estimate. Further work characterizing the heterogeneities of the distribution of serial intervals (11) and measuring serial intervals from individuals who are asymptomatic may help to improve the estimation of *R*_0_.

We calculated the herd immunity thresholds to be between 73% and 84% in China (6), the US and the eight European countries, based on the formula 1-1/*R*_0_ in a well-mixed population (39). Heterogeneity in population structure may lead to certain degree of deviations (42). Nonetheless, these are very high thresholds to reach even with an effective vaccine. For example, for an effective vaccine with 90% efficacy, a coverage of 90% of the population only leads to a population immunity of 81%. A recent survey showed that only approximately 50% of Americans plan to get a COVID-19 vaccine (34). Our results highlight the importance of public education about COVID-19 vaccination to ensure high vaccine coverage to achieve herd immunity (34).

It is not known how long the vaccine induced protective immunity lasts. We found that if the duration of immunity is relatively short as suggested in refs. (18, 19), and similar to the durations of protective immunity against other endemic coronaviruses (9, 20) or MERS-CoV (24), a frequent vaccination schedule once every couple of years to multiple times per year is needed to maintain herd immunity. However, a recent report showed that the T cell immunity to SARS-CoV-1 can last for 17 years in patients recovered from SARS-CoV-1 infection (22). In this case, a less frequent vaccination schedule is needed. Furthermore, we found that in addition to the mean duration of vaccine-induced protective immunity, the distribution of the duration is an important factor in determining vaccination frequency. A vaccine that induces a more uniform response in a population is better than a vaccine that induces a heterogeneous response in maintaining population immunity. Studies of the kinetics of antibody dynamics in individuals, such as refs. (19, 43), will help making more precise predictions of vaccine schedules. The stark difference between the durations of the protective antibody immunity and the T cell immunity (22) suggests that quantifying the individual heterogeneities in the kinetics of the T cell response will also be important.

Overall, our work shows that SARS-CoV-2 has high *R*_0_ values and spread very rapidly in the absence of strong control measures across different countries. This implies very high herd immunity thresholds, and thus highly effective vaccines with high levels of population coverage will be needed to prevent sustained transmission. If the protective immunity induced by vaccination is not long lasting, understanding the full distribution of the duration of protective immunity in the population is crucial to determine the frequency of vaccinations.

## Data Availability

All data is available in the main text and supplementary materials.

## Acknowledgments

We would like to thank Alan Perelson for suggestions and critical reading of the manuscript. The work is partially funded by the Laboratory Directed Research and Development (LDRD) Rapid Response Program through the Center for Nonlinear Studies at Los Alamos National Laboratory. RK and SS would like to acknowledge funding from DARPA (HR0011938513) and the Center for Nonlinear Studies. ERS was funded through the NIH grant (R01AI135946).

## Author contributions

RK and NH conceived the project; RK performed literature search and designed the study; SS collected data; RK, SS and ERS performed analyses; RK, SS, ERS and NH wrote and edited the manuscript.

